# Autozygosity mapping and time-to-spontaneous delivery in Norwegian parent-offspring trios

**DOI:** 10.1101/2020.06.25.20140103

**Authors:** Pol Sole-Navais, Jonas Bacelis, Øyvind Helgeland, Dominika Modzelewska, Marc Vaudel, Christopher Flatley, Ole Andreassen, Pål R. Njølstad, Louis J. Muglia, Stefan Johansson, Ge Zhang, Bo Jacobsson

**Affiliations:** Department of Obstetrics and Gynecology, Institute of Clinical Sciences, Sahlgrenska Academy, University of Gothenburg, Gothenburg, Sweden; KG Jebsen Center for Diabetes Research, Department of Clinical Science, University of Bergen, Bergen, Norway; Division of Health Data and Digitalization, Department of Genetics and Bioinformatics, Norwegian Institute of Public Health, Oslo, Norway; Center for Medical Genetics and Molecular Medicine, Haukeland University Hospital, Bergen, Norway; NORMENT, University of Oslo, Oslo, Norway; Division of Mental Health and Addiction, Oslo University Hospital, Oslo, Norway; Department of Psychiatry, University of California, San Diego, La Jolla, California; Department of Pediatrics and Adolescents, Haukeland University Hospital, Bergen, Norway; Department of Pediatrics, University of Cincinnati College of Medicine, Cincinnati, OH, United States; Division of Human Genetics, The Center for Prevention of Preterm Birth, Perinatal Institute, March of Dimes Prematurity Research Center Ohio Collaborative, Cincinnati Children’s Hospital Medical Center, Cincinnati, OH, United States; Department of Obstetrics and Gynecology, Sahlgrenska University Hospital, Gothenburg, Sweden

**Keywords:** preterm delivery, runs of homozygosity, genetics, Moba, pregnancy

## Abstract

Parental genetic relatedness may lead to adverse health and fitness outcomes in the offspring. However, the degree to which it affects human delivery timing is unknown. We use genotype data from ≃25,000 parent-offspring trios from the Norwegian Mother, Father and Child Cohort Study to optimize runs of homozygosity (ROH) calling by maximising the correlation between parental genetic relatedness and offspring ROHs. We then estimate the effect of maternal, paternal, and fetal autozygosity and that of autozygosity mapping (common segments and gene burden test) on the timing of spontaneous onset of delivery. The correlation between offspring ROH using a variety of parameters and parental genetic relatedness ranged between −0.2 and 0.6, revealing the importance of the minimum number of genetic variants included in a ROH and the use of genetic distance. The optimized parameters led to a ≃45% increase in the correlation between parental genetic relatedness and offspring ROH compared to using predefined parameters. We found no evidence of an effect of maternal, paternal nor fetal overall autozygosity on spontaneous delivery timing. Yet, using autozygosity mapping for common and rare autozygous segments, we identified three maternal *loci* in TBC1D1, SIGLECs and EDN1 gene regions reducing median time-to-spontaneous onset of delivery by ≃2-5% (p-value< 2.3×10^−6^). We also found suggestive evidence of a fetal *locus* at *3q22*.*2*, in the RYK gene region (p-value= 6.5×10^−6^). Autozygosity mapping may provide new insights on the genetic determinants and architecture of delivery timing beyond traditional genome-wide association studies, but particular and rigorous attention should be given to ROH calling parameter selection.

**Author summary:** Mating between relatives has an effect on offspring’s health and fitness in a number of species. In the offspring of genetically related parents, this is translated into long segments of the genome in the homozygous form (the same copy is inherited from each parent), but there is no consensus on how long these segments must be. In this study, we used dense genetic data from parent-offspring trios to optimize the detection of long segments of the genome. Our optimized long homozygous segments increased the correlation between parental genetic relatedness and offspring runs of homozygosity by ≃45% compared to widely used parameters. Furthermore, while preterm delivery is the global leading cause of mortality in children under 5 years, the degree to which long homozygous segments affect human delivery timing is unknown. We observed no maternal, paternal nor fetal effects of the proportion of the genome covered by homozygous segments on time-to-spontaneous delivery. However, by mapping these segments to the genome, we found evidence supporting three specific maternal segments falling on TBC1D1, SIGLECs and EDN2 gene regions to be associated with lower time-to-spontaneous onset of delivery. Future studies should assess the functional impact of these genes on spontaneous onset of delivery.

## Introduction

Offspring of genetically related parents may receive two copies of the same allele co-inherited from a common ancestor. Because alleles are likely to be inherited in long segments, the length of the genome covered by homozygous by descent alleles (autozygosity) rises. Yet, in each generation, recombination breaks autozygous segments into smaller ones, which will be subject to selection for a longer period of time. Consequently, long autozygous segments, a product of recent parental relatedness, are enriched in low-frequency and rare deleterious homozygous variants (1,2). The effects of parental genetic relatedness on general health and fitness (3) have been observed both in plants (4,5) and animals (6,7), including humans (e.g., reproductive success, overall health, height, lung function, and fluid intelligence (8–10)). The mapping of autozygous segments has provided insights into recessive effects in particular regions of the genome (11–14) by capturing the effects of low-frequency and rare non-genotyped damaging variants that lie within these segments (15), even in traits not affected by overall autozygosity (12,14,16).

Autozygous segments are generally detected by identifying segments of consecutive homozygous variants (runs-of-homozygosity, ROH) in dense genotype data (17–19). However, ROH may contain both homozygous alleles co-inherited from a common ancestor and non-autozygous alleles. The identification of truly autozygous segments requires the specification of various parameters (e.g., minimum ROH length, number of SNPs included), which, despite being optimized to detect autozygosity (17,20), remain arbitrary. These methodologic differences make between-study comparisons difficult and impede proper inference drawing.

Being the leading cause of death among children under five years of age (21), preterm delivery has a heavy burden on global health. The physiological control of human delivery timing is poorly understood; no adequate animal models exists, thus providing limited information about delivery timing mechanisms. Shaped by the maternal and fetal genomes, which are distinct, but otherwise related genomes, delivery timing is a trait with relatively high heritability (∼20-25% (22,23)). Yet, genome-wide association studies (GWAS) have had only limited success in discovering its genetic determinants, with some exceptions (23–25).

Here, we optimize ROH calling parameters by using the relationship between parental genetic relatedness and offspring ROH in parent-offspring triads and estimate the effect of maternal, paternal, and fetal autozygosity on the timing of delivery. We also adopted autozygosity mapping as an alternative to typical GWAS to identify segments of the genome potentially harboring low-frequency and rare genetic variants with recessive effects on spontaneous delivery timing.

## Results

### Parental genetic relatedness and offspring ROH

Before ROH calling in all triad members, we estimated parental genetic relatedness, and selected the parameters (LD pruning, physical vs genetic distance, minimum number of genetic variants included in a segment and the allowance of heterozygous calls within the segment) that maximized the correlation coefficient between parental genetic relatedness and offspring ROH, using 108 different combinations of parameters. We observed a limited amount of parental genetic relatedness, as expected in an outbred population, with a median of 5 cM shared between parents (Q1, Q3: 2.2, 12.4 total cM) (S1 Table).

The correlation coefficient between parental genetic relatedness and offspring ROH varied considerably depending on the sub-cohort and the parameters used to call ROHs in the offspring, ranging from −0.23 to 0.60 (S1 Figures). To understand the effect of the different parameters for each ROH call on the correlation coefficient, we ran a linear model using information from all cohorts. The minimum number of genetic variants to call a ROH (including a 3^rd^ degree polynomial) was the parameter with the strongest effect on the correlation coefficient, followed by the use of genetic vs physical distance and LD pruning (Figure 1A). Among the parameters maximizing the correlation coefficient, the use of genetic, as opposed to physical distance, was the only consistent parameter in all sub-cohorts (Figure 1B). Details on the optimal parameters used for each sub-cohort can be found in S2 Table, and the correlations between parental genetic relatedness and offspring ROH, using the optimized parameters can be visualized in Figure 2. The correlation coefficient between the optimized parameters and parental genetic relatedness was R≃ 0.55 (min R= 0.5; max R= 0.6), depending on the sub-cohort.

**Figure 1A.**
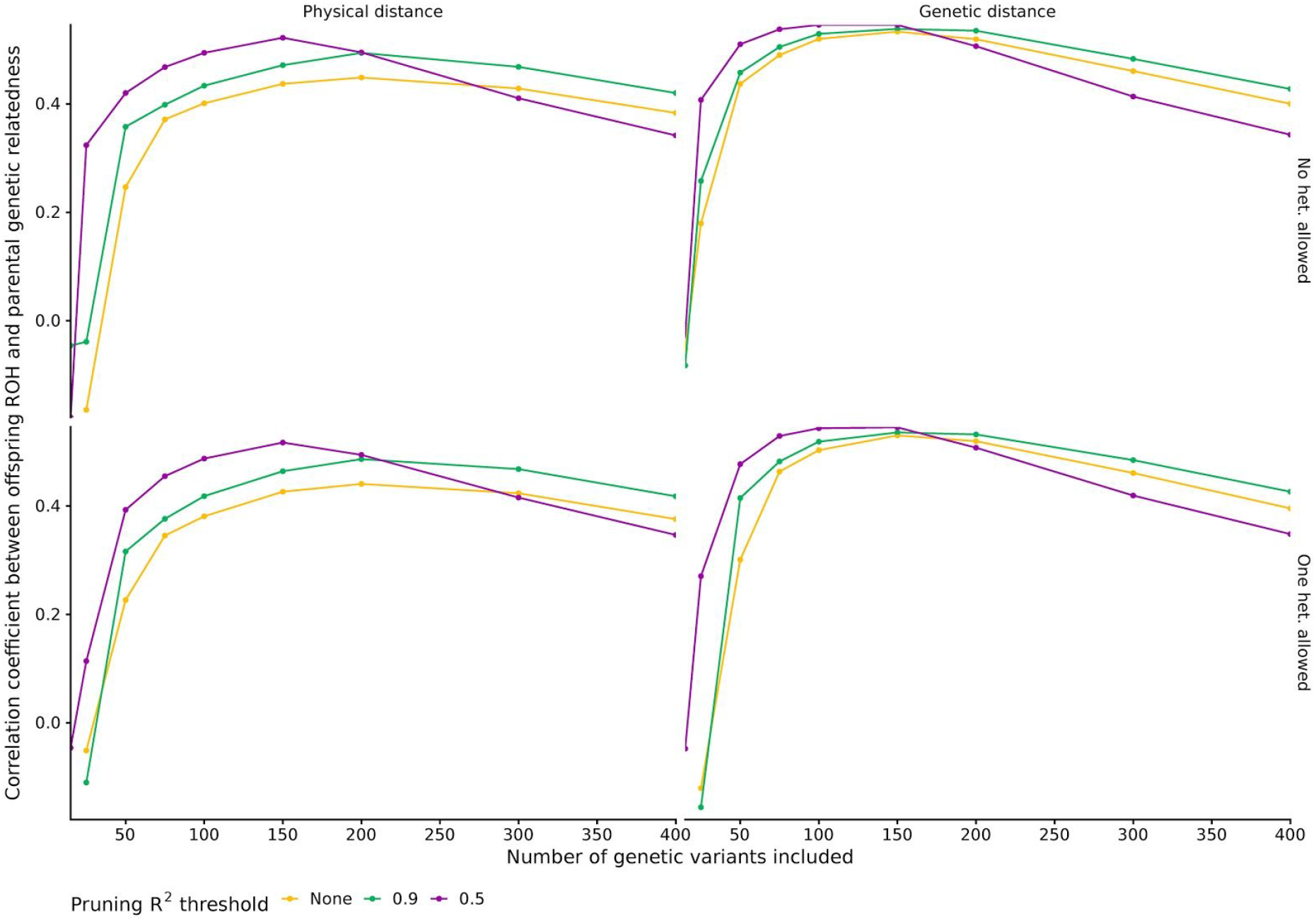
Minimum number of homozygous genetic variants included in a ROH segment and correlation coefficient between parental genetic relatedness and offspring ROH. The correlation coefficient was averaged across all sub-cohorts for visualization purposes (n= 24,927). Upper and bottom rows show the correlation coefficient when no or one heterozygous call was allowed, respectively. Left and right columns show the correlation coefficient when using physical or genetic distance, respectively.

**Figure 1B.**
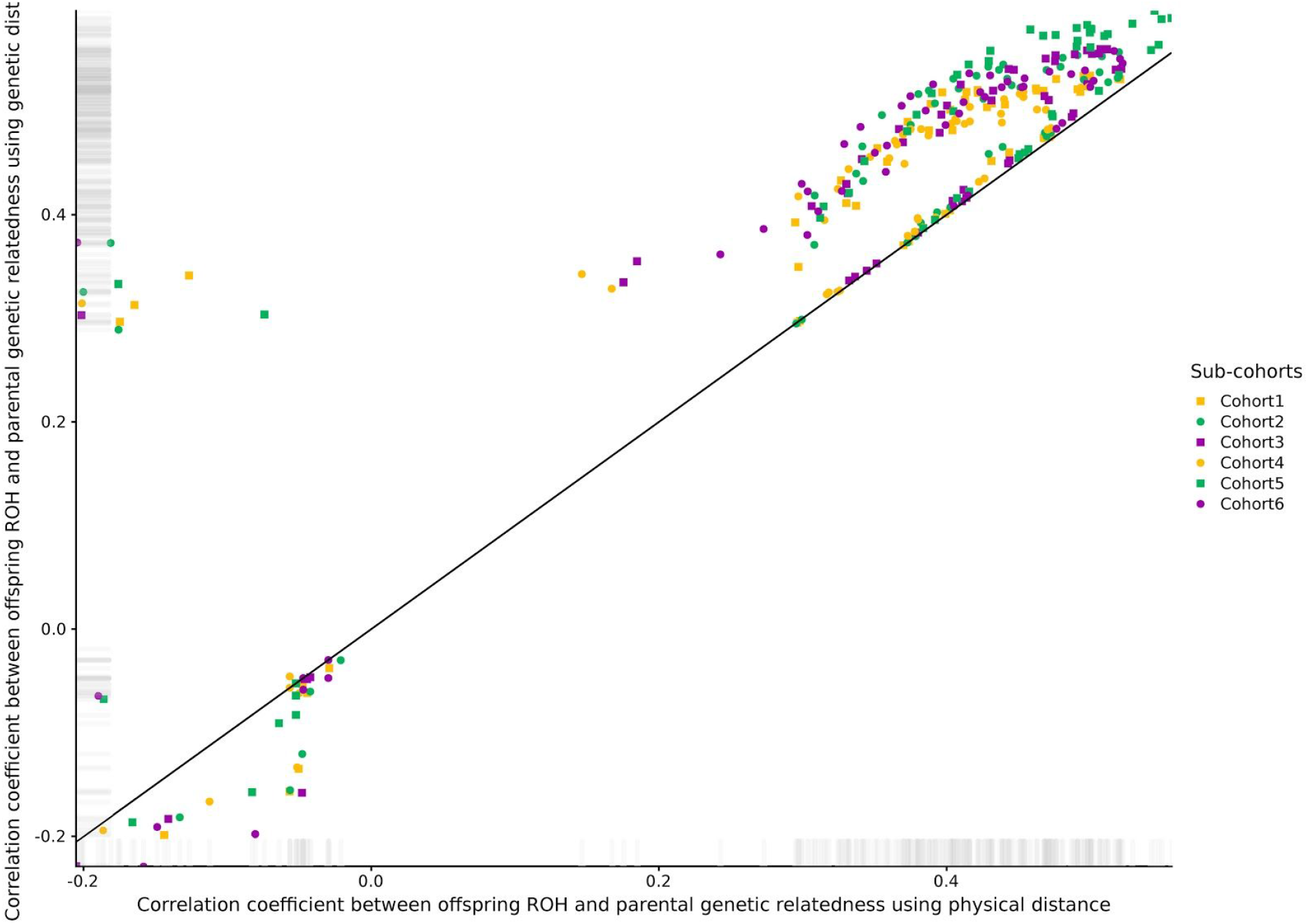
Correlation coefficient between parental genetic relatedness and offspring ROH using physical vs genetic distance. The correlation coefficient was averaged across all sub-cohorts for visualization purposes (n= 24,927). A total of 108 ROH calls were performed in offspring using different combinations of pruning, physical vs genetic distance, number of homozygous genetic variants and allowing one or no heterozygous calls within ROHs.

**Figure 2.**
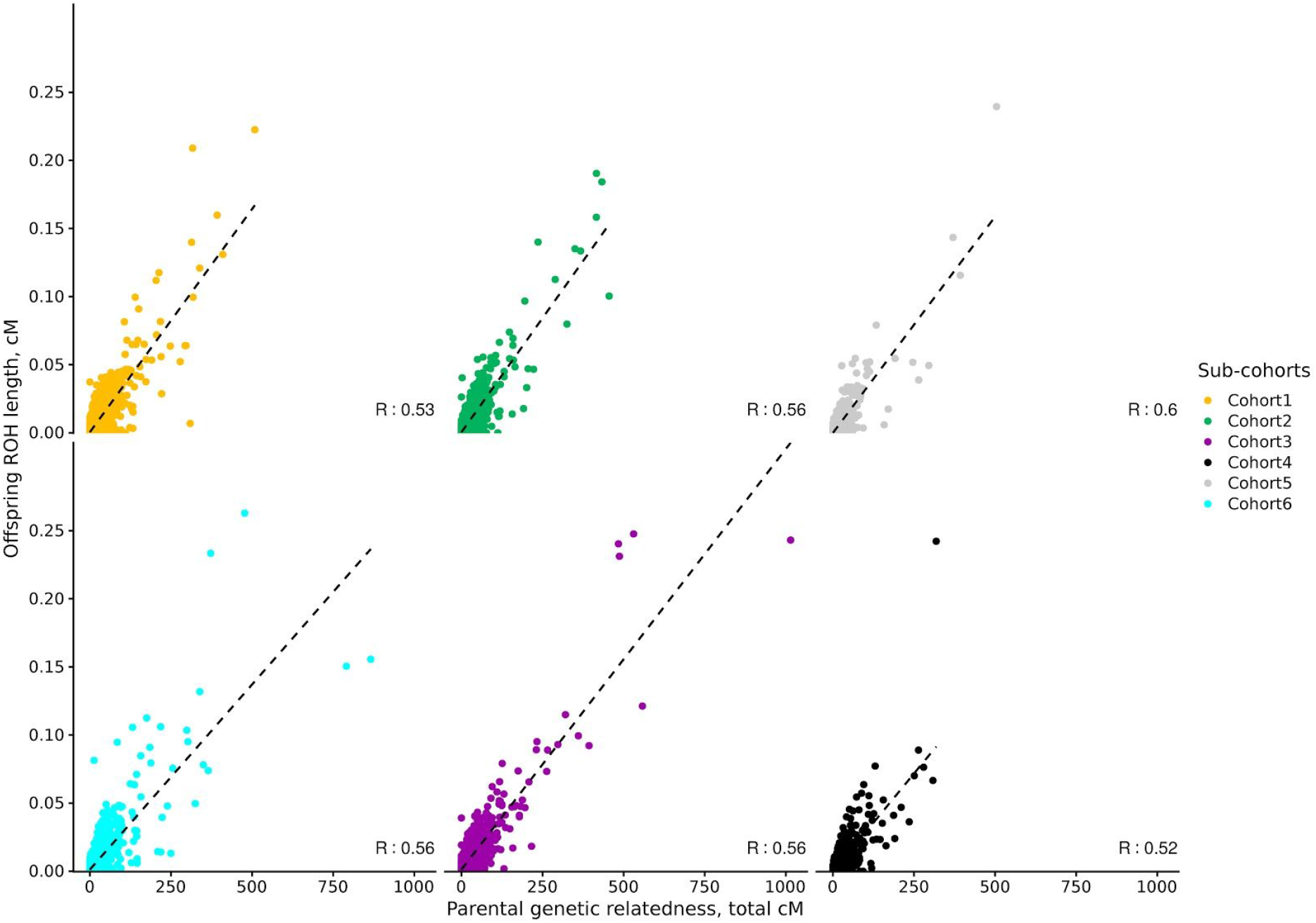
Offspring ROH and parental genetic relatedness using optimized ROH calling parameters. Total accumulated length of offspring ROH and parental genetic relatedness are in cM (n= 24,927). For each sub-cohort, ROH calling parameters explaining the highest proportion of parental genetic relatedness were selected, and are thus sub-cohort dependent.

### Estimated autozygosity in parent-offspring trios

Once the optimal parameters were identified for each sub-cohort (S2 Table), we called ROHs in all triad members using these optimized parameters. We sought to describe the distribution of F_ROH_ and the other measures of autozygosity between triad members (S3 Tables). We identified 15 mothers, 13 fathers and 9 fetuses with extreme inbreeding (F_ROH_ > 8%, proposed by Yengo L et al. (26)). ROHs were identified in 63.6% of the mothers (n= 23,676), 63.0% of the fathers (n= 24,805) and 57.2% of the fetuses (n= 23,948). We then compared the medians of different measures of autozygosity only in subjects with identified ROHs. F_ROH_, the number of autozygous segments and the average autozygous segment length (Figure 3) were lower in offspring than in both parents (Wilcoxon test, all p-values< 10^−16^), but we observed no differences between parents (for these analyses we did not include maternal chromosome X). As already described by Nalls et al. (27), a decline in measures of autozygosity after each generation is expected with increasing globalization and urbanization. Similarly, F_HOM_ was higher in both parents than in the offspring (median [Q1, Q3]: 0.0036 [-0.0009, 0.0079]), but was slightly higher in mothers (median [Q1, Q3]: 0.0046 [0.0001, 0.0092]) than in fathers (median [Q1, Q3]: 0.0039 [-0.0004, 0.0087]). Median time to most recent common ancestor was 16.1 generations in the mothers (Q1, Q3: 10.5, 23.0), 16.1 generations in the fathers (Q1, Q3: 10.4, 22.9) and 17.7 generations in the offspring (Q1, Q3: 12.0, 24.6).

**Figure 3.**
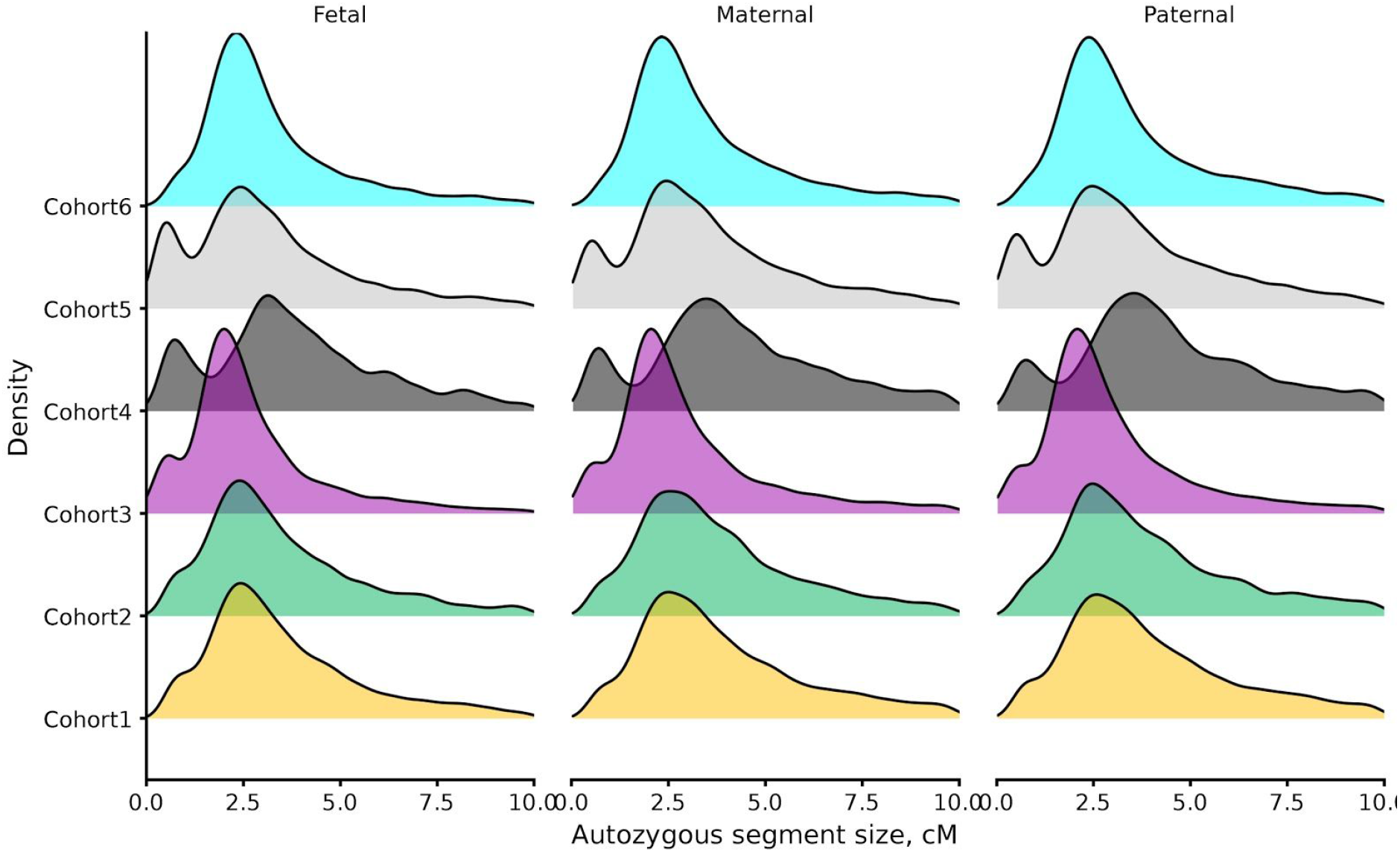
Distribution of the average segment length for each sub-cohort in maternal, paternal and fetal samples. Only subjects with detected autozygous segments are shown.

### Estimated autozygosity and delivery timing

To evaluate the effect of autozygosity on spontaneous delivery timing, we ran AFT models in mothers, offspring and fathers, using different measures of autozygosity (S3 Table). We observed no significant association between maternal (estimate: −0.06%; 95% CI: −0.17, 0.05%; p-value= 0.283, n= 21815, events= 19690), paternal (estimate: 0.09%; 95% CI: −0.02, 0.19%; p-value= 0.110 n= 21465, events= 19338) nor fetal F_ROH_ (estimate: −0.10%; 95% CI: −0.27, 0.06%; p-value= 0.216, n= 23503, events= 20789) and spontaneous delivery risk, in any of the models tested. The results obtained using F_HOM_, as well as the other measures of autozygosity, supported no effect of autozygosity on spontaneous delivery risk.

### Autozygosity mapping

We next investigated the frequency and effects of autozygous segments on spontaneous onset of delivery (S2 Figure). For each triad member group, we split overlapping segments from all sub-cohorts into unique segments. Most autozygous segments had a very low frequency, with ∼98% having a frequency below 1% (S2 Figure). All segments with a frequency >1% were identified in the HLA region (6p21), followed by segments in the LCT gene region (2q21.3), a region with recent positive selection (28), with some segments reaching a frequency of 0.8%.

For each of the identified autozygous segments, we ran an AFT model on spontaneous onset of delivery risk (S3 Figure). We obtained estimates for 60,391 (59,113 excluding chromosome 23) maternal, 58,742 paternal and 44,741 fetal segments. We identified 2 independent maternal segments (10 segments in total): one surviving a strict Bonferroni correction and the other surviving after correcting for the effective number of segments (p-value < 8.3×10^−7^ and 3.6×10^−6^, respectively). The high confidence loci is located in the TBC1D1 gene region (*4p14*). This segment, of 0.24 cM, had a frequency of ∼0.3% and was associated with a 2% lower median time-to-spontaneous onset of delivery (p-value= 7.4×10^−7^; Figure 4 and 5, Table 1 and S4 Figure). TBC1D1 encodes the TBC1 Domain Family Member 1 protein, which has a role in regulating cell growth and differentiation. TBC1D1 is highly expressed in female reproductive organs (cervix, uterus and vagina (29)), but genetic variants affecting TBC1D1 expression in these organs are yet to be identified. Nonetheless, if we consider non-reproductive organs, a number of eQTLs affecting TBC1D1 expression have been identified in thyroid glands, with some observational studies showing an association between thyroid function and preterm delivery (30,31).

**Table 1.** Independent high and low confidence autozygous segments associated with time-to-spontaneous onset of delivery.

| Chr. | Start | End | Nearest gene | N | Freq. | Effect size (%)<sup>a</sup> | P-value | Mim |
| --- | --- | --- | --- | --- | --- | --- | --- | --- |
| **Maternal** |  |  |  |  |  |  |  |  |
| 4 | 3,788,4116 | 3,790,0371 | TBC1D1 | 23,323 | 0.003 | -2.1 | 7.4x10<sup>-7</sup> | - |
| 4 | 3,758,6331 | 3,759,0112 | c4orf19 | 23,323 | 0.002 | -2.1 | 2.3x10<sup>-6</sup> | - |
| **Paternal** |  |  |  |  |  |  |  |  |
| 12 | 7,608,6784 | 76099541 | PGAM5 | 22,901 | 0.003 | -1.9 | 3.2x10<sup>-6</sup> | - |
| **Fetal** |  |  |  |  |  |  |  |  |
| 3 | 13,398,9068 | 13,4013,144 | SPSB4 | 23,332 | 0.001 | -2.7 | 2.0x10<sup>-6</sup> | - |
| 3 | 13,401,3144 | 13,403,3585 | ARL14 | 23,332 | 0.001 | -2.7 | 2.0x10<sup>-6</sup> | - |

High and low confidence autozygous segments (p-values below Bonferroni correction for effective number of segments) are shown. For each segment, the closest protein-coding gene is also shown, and for this gene, the Mim phenotype number for a phenotype with recessive inheritance. The total number of segments (effective number of segments) was 59,193 (13,803), 58,742 (13,328) and 44,741 (11,982) for maternal, paternal and fetal samples, respectively.
<sup>a</sup> The effect size reflects the effect of the autozygous segment on time-to-spontaneous onset of delivery, representing the percent difference in time-to-spontaneous onset of delivery between subjects affected by an autozygous segment in the region vs those not affected.

**Figure 4.**
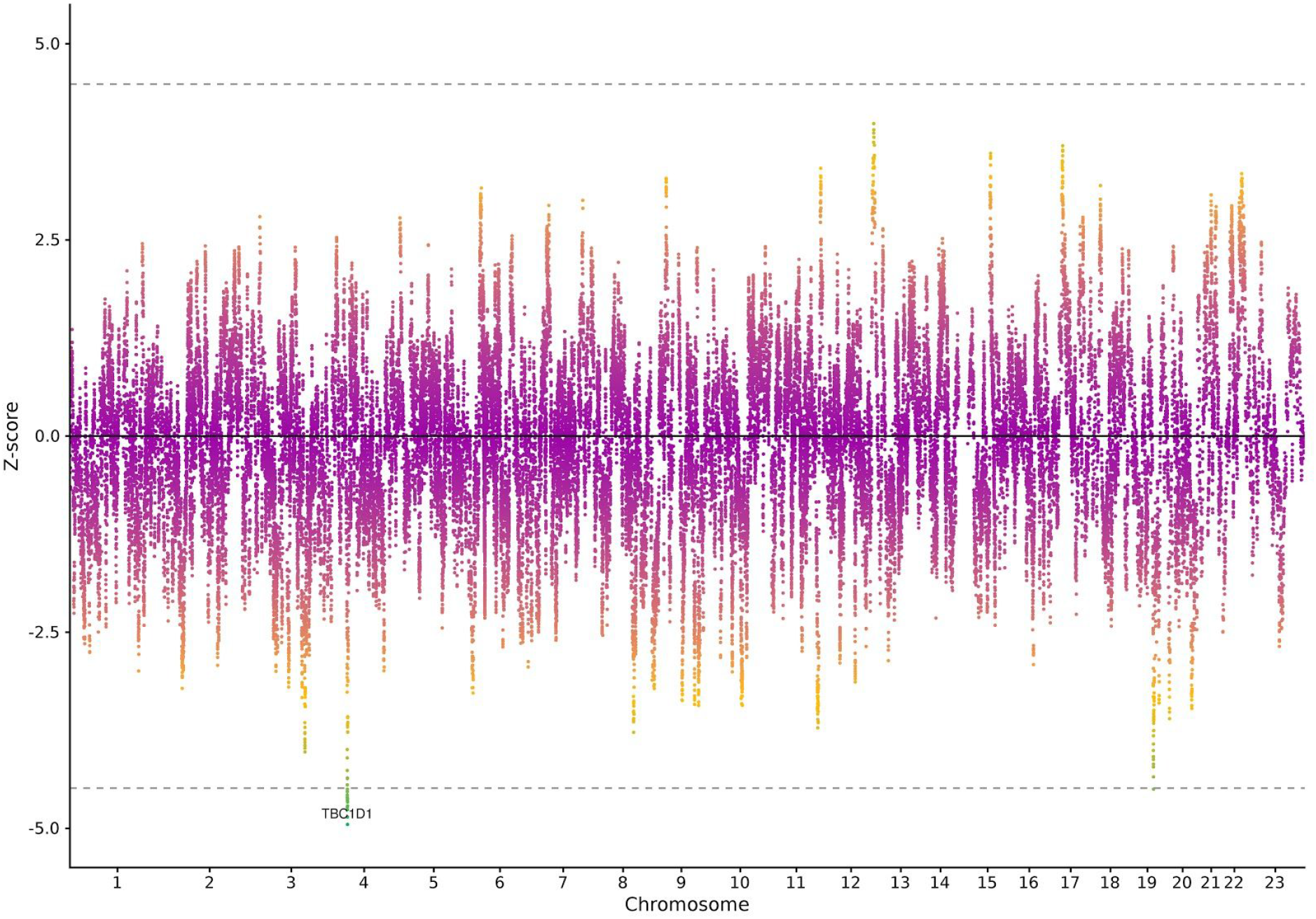
Associations between maternal autozygous segments and time-to-spontaneous delivery. Z-scores of maternal autozygous segments obtained from accelerated failure time models on time-to-spontaneous onset of delivery. The Bonferroni threshold for significance (n= 23,323, n autozygous segments= 60,391, effective n of segments= 13,803) is indicated by the dotted line.

**Figure 5.**
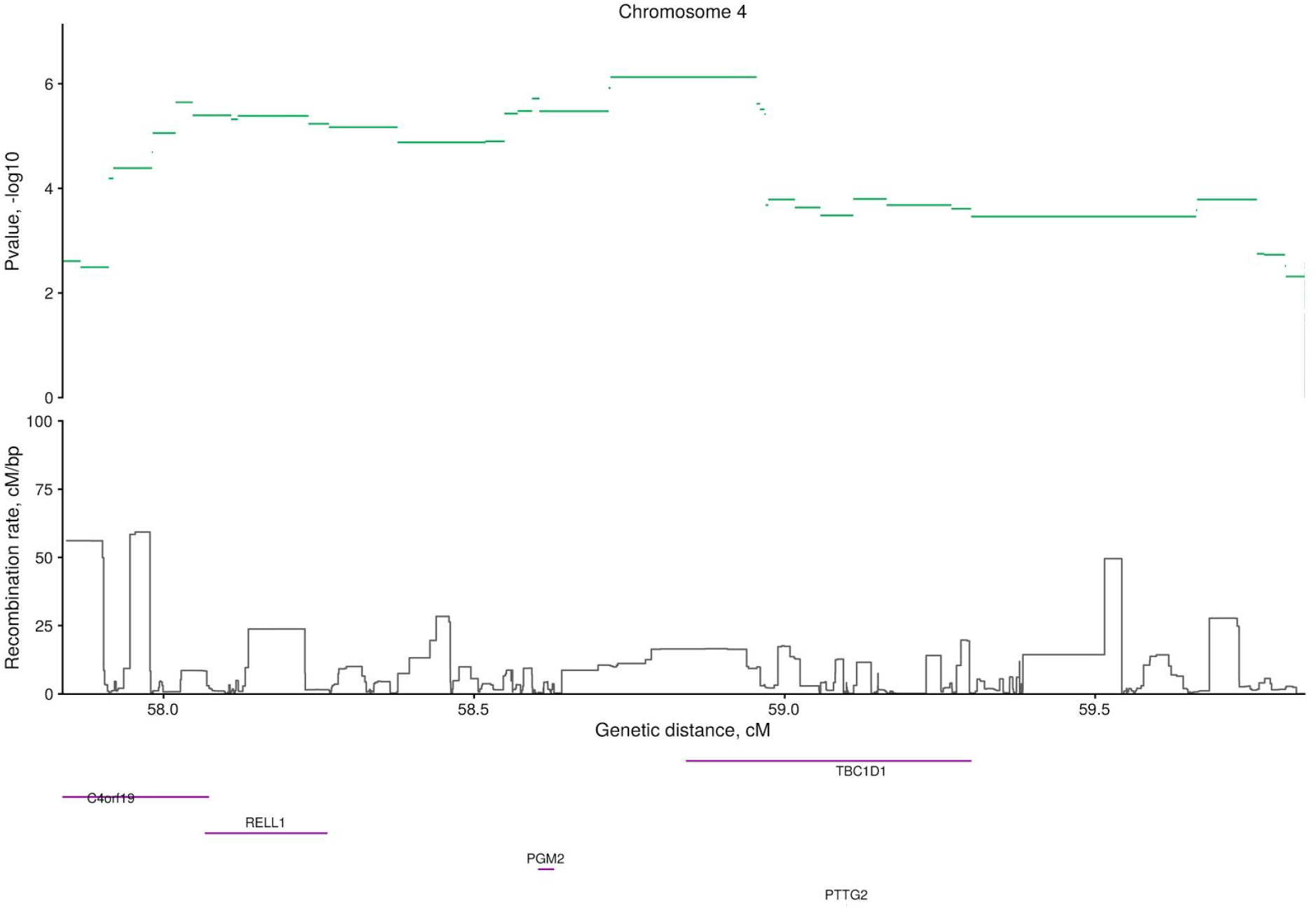
Associations between autozygous segments at the maternal *4p14 locus with* time-to-spontaneous onset of delivery. P-values were obtained from accelerated failure time models on time-to-spontaneous onset of delivery (n= 23,323). Gene names for the ten longest genes in the region and recombination rate are also depicted.

**Figure 6.**
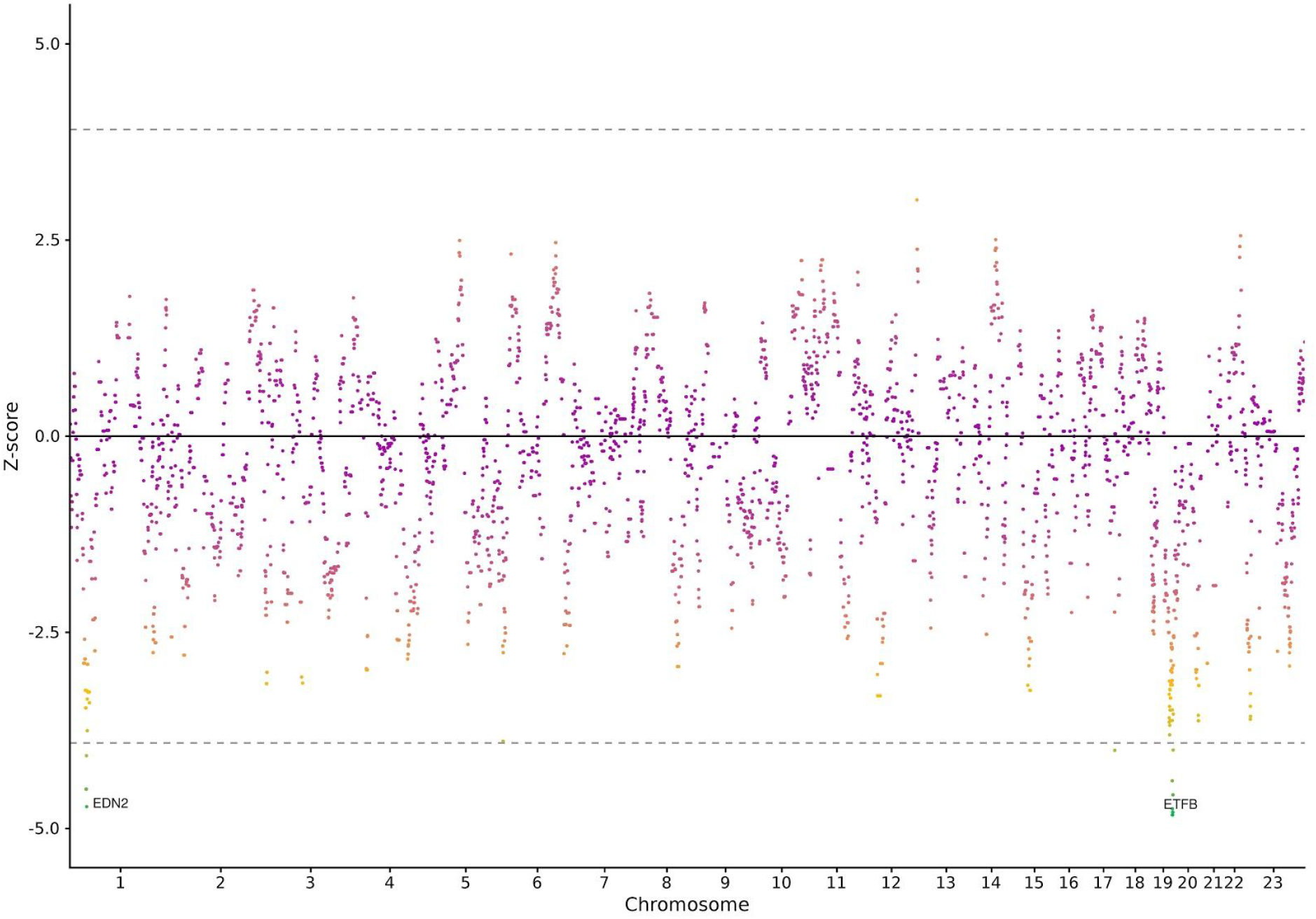
Maternal long segment gene burden test and time-to-spontaneous delivery. Z-scores of maternal genes obtained from accelerated failure time models on time-to-spontaneous onset of delivery are shown. Long autozygous segments were collapsed into protein-coding genes as a binary variable. The dotted line indicates the Bonferroni threshold for significance correcting for the effective number of genes (n= 23323, n genes= 18675, effective n of genes= 1084).

We repeated the analysis in ∼2,500 mothers, using predefined parameters (see Materials and methods), but we were unable to detect autozygous segments overlapping any of the high or low confidence segments.

We didn’t identify any high confidence segment in neither fathers nor fetuses (S5 Figures). Yet, using the effective number of autozygous segments (13,328 and 11,982, respectively for fathers and fetuses), we obtained suggestive evidence for an additional independent *locus* in fathers and two independent *loci* in fetuses. The fetal *loci* were located in chromosome 3, near SPSB4 and ARL14 gene regions. The two segments, with a frequency of 0.01%, reduced the time-to-spontaneous onset of delivery by 2.7%. While the center of each segment is >1 cM apart from each other, the two fetal *loci* have the same effect size, standard error and frequency, suggesting that these are not independent. SPSB4 encodes a protein involved in innate immune system and ARL14 controls the major histocompatibility complex I transport in dendritic cells. The paternal low confidence autozygous segment was observed in a non-coding region, had a frequency of 0.03%, and an effect on time-to-spontaneous onset of delivery of -2%.

We did not expect an effect of paternal autozygous segments, and therefore viewed autozygosity mapping in fathers as a negative control to detect bias due to uncontrolled confounding (e.g., population stratification). We discovered a paternal *locus* only after correcting for the effective number of segments. While this provides a substantial degree of assurance regarding the robustness of the maternal high confidence segment identified, the same cannot be argued for low confidence segments in neither mothers, fathers, nor fetuses. This was confirmed by Q-Q plots of p-values (S3 Figure). As seen in rare variant analysis, this might be due to the overall low frequency of autozygous segments which are a more recent event, and may tend to be geographically clustered (32).

### Survival analysis of imputed data under a recessive model

Before running AFT models using single genetic variants, we attempted to identify homozygous subjects (mothers, fathers and fetuses) for imputed genetic variants within the high and low confidence segments with protein consequences. GNOMAD v2.1.1 reports no genetic variants with a predicted consequence within the top maternal segment (TBC1D1 gene region), or for any of the low confidence segments in fathers nor fetuses. We identified none but 1 mother homozygous for two missense variants (rs187076049 and rs10001580) within the phosphoglucomutase 2 (PGM2) gene region. This subject had a gestational duration below the median, and its removal slightly attenuated the effect size and increased the standard error, resulting in an increase in the p-value (from 1.9×10^−6^ including the subject to 4.1×10^−6^ after its removal). The rs10001580 genetic variant is associated with oestradiol levels (33), and is an eQTL for PGM2 in cultured fibroblasts (29) and blood (34), and for TBC1D1 in blood (34).

Intending to identify genetic variants underlying the effects observed in autozygosity mapping, we performed survival analysis under a recessive model using imputed genetic data for all variants within the high and low confidence segments (S5 Table). Results included 580 maternal, 65 paternal, and 164 offspring genetic variants. We identified an intronic maternal genetic variant in the PGM2 gene associated with time-to-spontaneous onset of delivery after genome-wide Bonferroni correction (rs76770307, p-value= 1.2×10^−9^), but with a very low homozygous count (n= 1). None of the two genetic variants reported above (rs187076049 and rs10001580) were associated with time-to-spontaneous onset of delivery using a recessive model (p-value= 0.431 and 0.828). Overall, 14 maternal genetic variants passed a relaxed Bonferroni correction (p-value< 0.05 / 580), with only 2 genetic variants with a homozygous count >5 (rs10029748, located 5’ UTR of TBC1D1 and rs10008243, an intronic variant in TBC1D1). We were unable to replicate these signals, given that, while imputed, these genetic variants had a homozygous count of 0 in the replication dataset.

We identified no paternal genetic variants associated with time-to-spontaneous onset of delivery, even using a relaxed Bonferroni correction (0.05/ 65).

Along the same lines, no fetal genetic variants had a genome-wide significant p-value, and only one genetic variant (rs77926300) passed after applying a relaxed Bonferroni correction (0.05/ 164). However, this genetic variant had a homozygous count of 1.

### Long segment gene burden analysis

Gene-level burden analysis of long autozygous segments provided evidence for additional genes (S6 Figure). We analysed a total of 18675 maternal, 17931 paternal and 17621 fetal protein-coding genes. After controlling for multiple comparisons, we identified 41 different genes in mothers (Figure 5) in the *19q13*.*41 locus* (p-values< 2.0×10^−6^), most probably as part of the same autozygous segment, and EDN2 at the *1p34*.*2 locus* (p-value= 2.3×10^−6^). The *19q13*.*41 locus* contains numerous Sialic acid-binding immunoglobulin-type lectins (SIGLECS) and zinc finger protein genes. SIGLECs are cell surface proteins that bind sialic acid, and are mostly expressed in immune cells. Some of the 14 SIGLEC genes that exist, are also expressed in villous and extravillous cytotrophoblasts, decidual cells, as early as 8 gestational weeks and in maternal uterine glands (35,36), suggesting a possible role of SIGLECS in mediating the immune tolerance at the feto-maternal interface. EDN2 gene (*1p34*.*2 locus*), associated with a almost 5% lower time-to-spontaneous onset of delivery (p-value= 2.3×10^−6^), is a strong vasoconstrictor involved in follicular rupture and ovulation; it causes the contraction of the smooth muscle layer surrounding each follicle (37,38). TBC1D1 gene, the top associated segment in autozygosity mapping, was significantly associated with time-to-spontaneous onset of delivery at the nominal level (p-value= 0.003) in the gene burden analysis.

Given the nature of this analysis targeting low frequency segments, we were unable to detect an effect of these genes in 2500 mothers.

We identified no paternal genes, and only one fetal gene, DAOA (*13q33*.*2* region) after controlling for multiple comparisons (S7 Figures).

### Comparison against predefined parameters

For completeness, we compared our approach with previously suggested (9) and widely used (10,26,39,40) parameters for ROH calling. Compared to the optimized parameters, predefined parameters led to an average 45% reduction in the correlation estimates between parental genetic relatedness and offspring ROH (Table 2 and S8 Figure). We then assessed, for each individual, the overlap between segments called using optimized parameters and those using Joshi’s parameters; the optimized parameters served as reference. We observed no overlapping segments in 29% of subjects, and an overlap >90% in 58% of subjects (Figure 7).

**Table 2.**
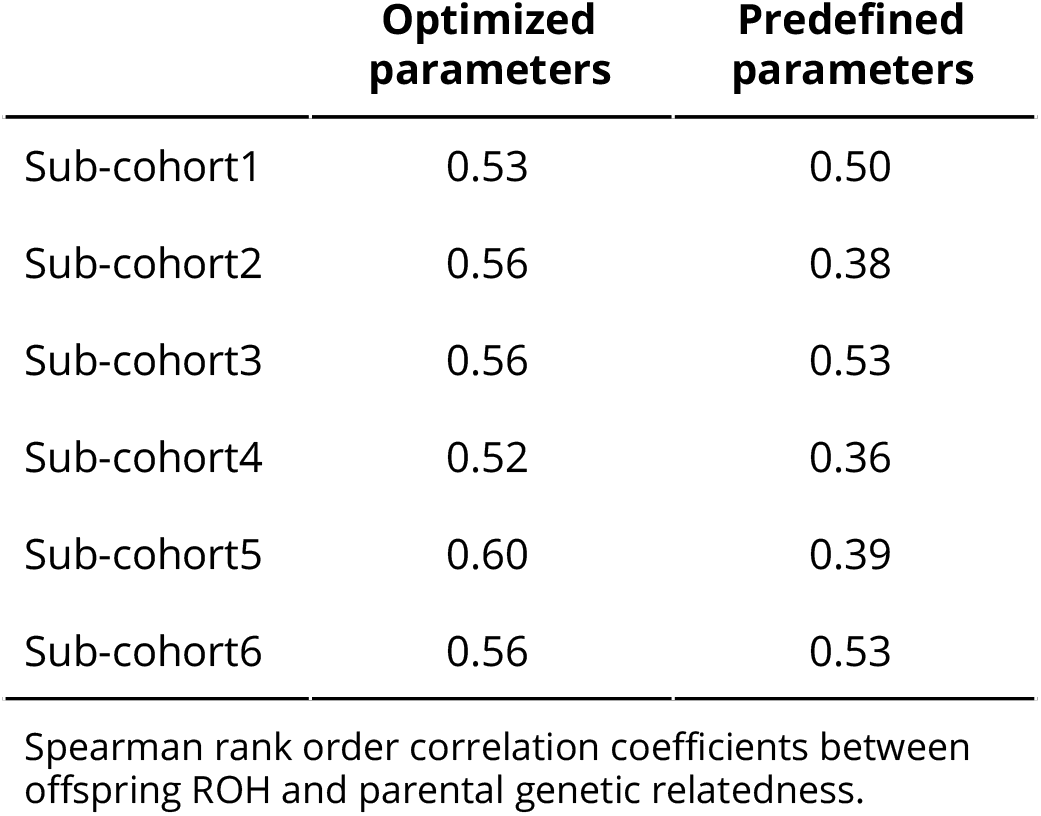
Correlation coefficients between offspring ROH and parental genetic relatedness using the optimized and predefined parameters.

|  | **Optimized**<br>**parameters** | **Predefined**<br>**parameters** |
| --- | --- | --- |
| Sub-cohort1 | 0.53 | 0.50 |
| Sub-cohort2 | 0.56 | 0.38 |
| Sub-cohort3 | 0.56 | 0.53 |
| Sub-cohort4 | 0.52 | 0.36 |
| Sub-cohort5 | 0.60 | 0.39 |
| Sub-cohort6 | 0.56 | 0.53 |

Spearman rank order correlation coefficients between offspring ROH and parental genetic relatedness.

**Figure 7.**
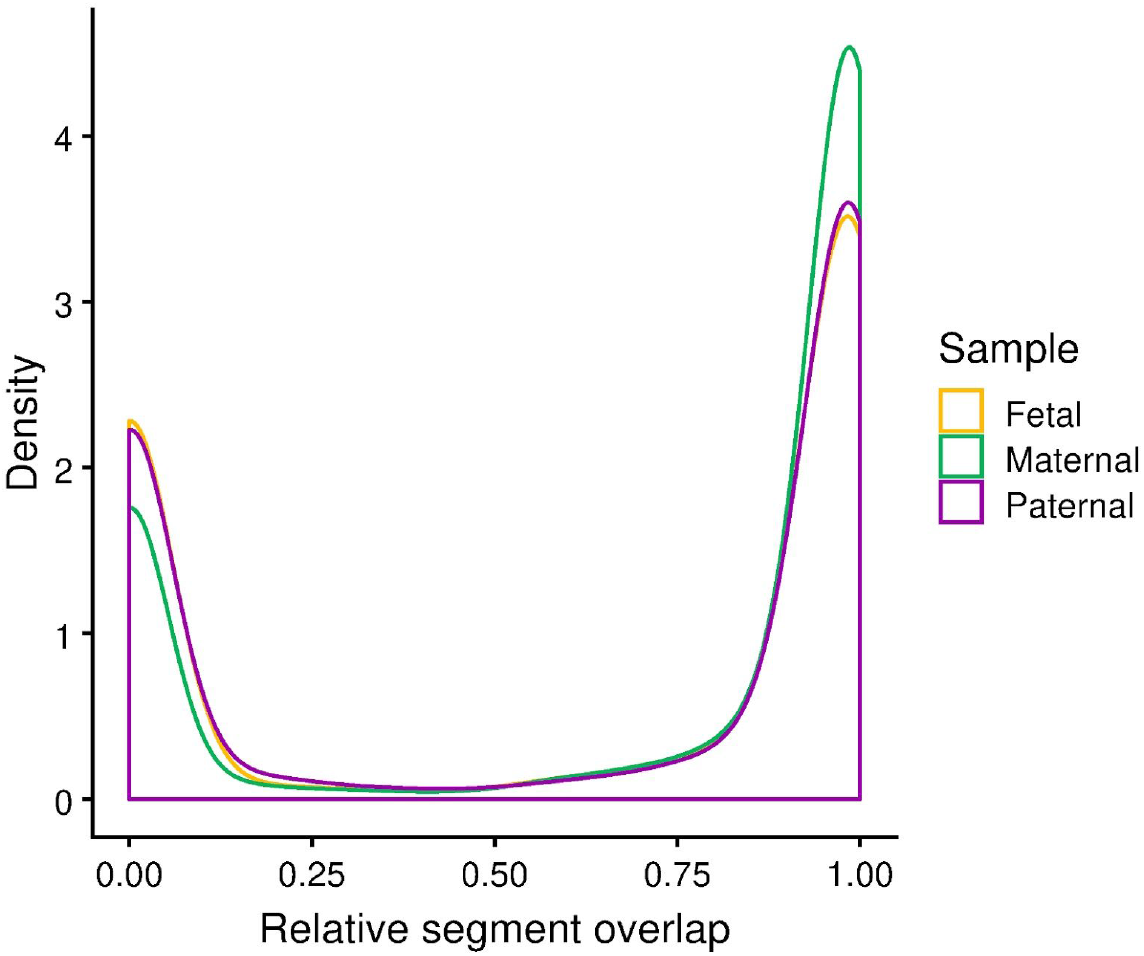
Autozygous segment overlap between optimized and predefined ROH parameters in mothers, fathers and fetuses. For each individual, we calculated the proportion of overlap between autozygous segments called using the optimized and predefined parameters. The optimized parameters served as the reference. Results for all cohorts merged are shown.

## Discussion

Low-frequency and rare genetic variants may play an essential role in human delivery timing, but individual variants are difficult to identify due to a lack of information on gestational duration in already available large sequencing datasets (e.g. UK Biobank). Here, we systematically called ROH segments using over 20,000 parent-offspring trios, and assessed the relationship between maternal, paternal and fetal autozygosity and spontaneous delivery risk. Calling ROHs using cohort-specific, rather than predefined, parameters increased the correlation between offspring ROH segments and parental genetic relatedness by 45%. While we observed no effect of overall autozygosity on delivery timing, we identified three maternal autozygous segments in *4p14, 19q13*.*41* and *1p34*.*2* loci associated with time-to-spontaneous onset of delivery. We found no supporting evidence suggesting that imputed genetic variants within these segments were driving the observed effects under a recessive mode.

ROH calling parameter selection based on the relationship between offspring ROH length and parental genetic relatedness provided us with non-arbitrary F_ROH_ estimates. As a consequence, ROH calling parameters varied for each sub-cohort, leading to differences in the average length and the total number of autozygous segments between sub-cohorts. While the optimized parameters were far from optimal (R ∼ 0.55), our results indicate that the use of predefined parameters would have performed worse (R ∼0.4). We observed this in samples genotyped with different arrays, but with a similar population structure; we expect differences to be larger in samples with more diverse population background. We sought to remove arbitrariness in the selection of parameters, yet it is unavoidable to use subjective parameters, for example, when estimating parental genetic relatedness with GERMLINE. The use of an alternative minimum length for IBD segment identification would have probably led to different optimized parameters. We selected a minimum length of 2 cM, for which GERMLINE has a detection power and accuracy of ∼70% (41). Another limitation was assuming that the optimized parameters we identified in the offspring would also be applicable to parents. While this assumption may not hold, a formal test would require parental and grand-parental genetic data, which is not available.

We observed no maternal, fetal nor paternal effect of autozygosity on time-to-spontaneous onset of delivery, either indicating there is no effect or the effect is particularly modest. Despite a consensus for the use of F_ROH_ when estimating autozygosity, we used four additional alternative measures: all confirming a null effect. Given the small amount of autozygosity detected in outbred populations, large sample sizes are required for the detection of effects on any trait. Consistent with our findings, two recent studies in 200 000 and 1M individuals found no effects of autozygosity on their own birth weight (8,10), a measure strongly correlated with the duration of gestation. We did not classify ROHs according to their length, which would have helped to understand recent parental relatedness, background relatedness, or founder events in our sample (42). However, using our approach we were targeting segments arising from recent parental genetic relatedness. Moreover, using 2 cM as the minimum threshold for IBD detection indicates that the common ancestor is 5-6 generations from parents (43).

IBD mapping offers better power than GWAS (44), which could be also true for autozygosity mapping. While we identified one region of high confidence in the TBC1D1 gene region, we find no evidence of association from imputed genetic variants within this region using a recessive model. The underlying causal genetic variant, if any, might be rare and in low LD with other variants, suggesting that it is likely to affect protein sequence rather than the expression. Autozygosity mapping is similar to GWAS in the sense that it does not provide evidence of causality: non-controlled population structure can also result in significant differences in ROH rates in relation to time-to-spontaneous onset of delivery. As expected, and supporting a robust population stratification control, we did not observe a paternal effect of autozygosity, nor of autozygosity mapping using a stringent Bonferroni correction. However, we admit that we identified one paternal autozygous segment associated with time-to-spontaneous onset of delivery when controlling for the number of effective segments. As such, we only considered the results obtained controlling for the total number of autozygous segments.

Genomic proximity does not always guarantee correct gene identification (45). By assuming that autozygous segments act through similar mechanisms to monogenic traits, the mapping of autozygous segments to causal genes may appear to be less difficult than in GWAS. However, we expressly targeted long segments, which span multiple genes, making correct gene identification diffcult. Hence, while the autozygous segment in TBC1D1 gene region was the top associated segment, segments in the same region overlapping with other genes (PGM2) were also highly significant. Maternal autozygous segments in TBC1D1 gene region were associated with 2.5% lower time-to-spontaneous onset of delivery. This gene, as well as PGM2, is highly expressed in the cervix, uterus and vagina, adding certain reassurance of a true positive. TBC1D1 gene expression in the endometrium of cattle is increased during pregnancy (46), and TBC1D1 is involved in muscle-contraction glucose uptake (47,48). While in this study we targeted autozygous segments resulting from recent parental genetic relatedness, we decided to target long runs of homozygosity and perform a gene burden test. This provided additional insights, involving two loci, which are also highly expressed in either placenta, uterus or ovaries: SEGLECs and EDN2. The effect size of these genes was twice as big as the effect size observed for the segments identified through autozygosity mapping. SIGLEC-6 plays a role in placental excess proliferation and invasion (49), and its expression is increased in the basal plate and chorionic villi of preterm preeclamptic mothers, but not in those delivering at term (35). EDN2 is a strong vasoconstrictor, and causes the contraction of the smooth muscle layer surrounding each follicle (37,38). EDN2 mediates the ovulation by inducing the contraction of follicles for oocyte expulsion (36,37). The expression of a MicroRNAs that targets EDN2 (miR-210) was decreased in placental villous tissues from preterm deliveries compared to term deliveries (50). Whether this gene can induce the contraction of other smooth muscles, such as endometrium, remains unknown. All these genes appear to be pointing towards either placental complications, (gestational) diabetes, or medical conditions linked to high blood pressure. However, in this study we explicitly excluded subjects with placental complications, diabetes or gestational diabetes, hypertension and preeclampsia, potentially ruling out a mediation of the observed effects through any of these conditions.

In the absence of large sequencing data with high-quality information on pregnancy phenotypes, moving beyond traditional GWAS may prove to be useful to identify *loci* associated with delivery timing. However, special attention must be put on ROH calling parameters, particularly in the absence of parent-offspring data. Our observations suggest that there is no or a very small effect of autozygosity on spontaneous delivery timing. Autozygosity mapping and gene burden tests highlighted three candidate maternal *loci* associated with time-to-spontaneous onset of delivery. We hope that future functional follow-up studies based on the observations presented here will yield novel insights and a better characterisation of the mechanisms behind human delivery timing.

## Materials and methods

### Study population

In this study, we used genotype data drawn from the Norwegian Mother, Father and Child Cohort study (MoBa) (51,52). This family-based cohort enrolled more than 114,000 children, 95,000 mothers, and 75,000 fathers from 50 Norwegian Hospitals between 1999 and 2008. The MoBa Genetics infrastructure is a collaborative research effort consisting mainly of three major research projects, with a total of ∼25,000 parent-offspring trios genotyped. Four different genotyping arrays were used in each project: 11,000 parent-offspring trios were genotyped with Illumina HumanCoreExome at the Genomics Core Facility (Norwegian University of Science and Technology, Trondheim, Norway), 9,000 parent-offspring trios were genotyped with Illumina Infinium Global Screening Array MD at the Erasmus Medical Center (Erasmus University, Rotterdam, the Netherlands), 3,000 parent-offspring trios were genotyped using Infinium Global Screening Array 24 at deCode Genetics (Reykjavik, Iceland) and 5,000 parent-offspring trios were genotyped using Illumina Infinium OmniExpress at deCode Genetics (Reykjavik, Iceland). For the replication of single genetic variant results, we obtained additional genetic data from 3,000 mothers from the same MoBa cohort, genotyped in two batches (Illumina Infinium OmniExpress 24, genotyped at deCode Genetics, Reykjavik, Iceland).

We excluded multiple pregnancies, women with gestational or type 2 diabetes, women with pre-eclampsia or hypertension, pregnancies lasting <154 (considered not viable) or ≥308 days, with a birth weight below 1,500 gr, missing gestational duration, conceived by in-vitro fertilization, or affected by any of the following: polyhydramnios, oligohydramnios, or congenital malformations. Gestational duration was estimated by ultrasound scan at 19-20 gestational weeks.

### Phenotype and covariates

Maternal health information prior to and during pregnancy, including gestational duration and parity, as well as complications of pregnancy and birth were recovered from the Medical Birth Registry of Norway (MBRN).

Descriptive characteristics of the different sub-cohorts can be viewed in S6 Table. We defined spontaneous onset of delivery as a delivery initiated by spontaneous contractions or rupture of membranes. Deliveries initiated by induction methods, including the use of prostaglandins, oxytocin, amniotomy or any other induction procedure, or planned cesarean section were censored. Maternal education was defined as ≤12 years, 13 to 16 years or ≥17 years, and household incomes as none, one parent or both of parents having an income >300,000 Norwegian crowns.

### Genotyping and quality control

Genotyping and quality control have been previously described (53), and performed accordingly for all samples included in this study, regardless of the genotyping array platform used. Samples with a call rate <0.98 or excess heterozygosity >4 SD, and variants with call rates <98%, 10% GenCall-score <0.3, cluster separation <0.4, Theta AA standard deviation >0.4, and HWE p-value <10^−6^ were excluded. We included autosomal markers for fathers and offspring, as well as autosomal chromosome X markers for mothers. Genome coordinates were mapped to the Genome Reference Consortium Human Build 37 (hg19). We excluded samples with recent ancestry different to European, and those with genetic relatedness with another sample greater than second to a third cousin (KING (54) cut-off> 0.0884). After ROH calling in mothers, fathers and offspring, we excluded subjects with suspected uniparental disomy defined as having at least one chromosome with >70% covered by ROH. Subjects with extreme inbreeding, defined as ROH segments covering >8% of the genome, were also excluded.

### Autozygosity parameter selection

ROH calling with PLINK (55) relies on the input of several parameters such as the minimum ROH length, the number of heterozygotes allowed, or the number of genetic variants included, amongst others. Despite the fact that these parameters have been optimized for estimating autozygosity (17,20), the selection remains arbitrary. To overcome this, we selected PLINK v.1.9 parameters *a posteriori*. We called ROH segments 108 times in the offspring using different parameters and selected the parameters that maximized the Spearman rank-order correlation coefficient between parental genetic relatedness and offspring ROH length. These parameters were later used to call ROHs in mothers, fathers, and offspring (see Autozygosity calling in parent-offspring trios). Parental genetic relatedness was estimated as the total length of shared identical-by-descent (IBD) segments in cM. Parental IBD was estimated using GERMLINE v.1.5.3 (43) (minimum length of IBD shared segments: 2 cM; remaining parameters set by default). Prior to IBD identification, haplotype phase was estimated using Eagle v.2.4.1 (56) using trio data. We filtered out variants with minor allele frequency <0.05 and pruned fetal genomic data using three different LD thresholds no, soft (R^2^ >0.9) and moderate (R^2^ >0.5) pruning. According to Gazal S. et al. (20), ROH calling using genetic distance may lower the variability of the estimate. However, PLINK v.1.9 does not allow ROH calling based on cM; we replaced the physical distance (base pairs) by the genetic distance (cM) extracted from 1000 Genomes Project (57). The conversion of physical to genetic distance of individual genetic variants was performed by linear interpolation. Due to PLINK bim file format not allowing base pairs to be recorded as decimals, we multiplied the genetic distance by 10^6^ to obtain a unique list of coordinates in cM within each chromosome. We called ROHs using PLINK’s --homozyg command 108 times, by modifying the LD pruning threshold, using cM or bp, varying the number of heterozygotes allowed and the minimum count to call a ROH. Similar to Howrigan et al. (17), we kept SNP density (--homozyg-density, high to ignore: 5000 kcM or kb), kb or cM minimum distance (--homozyg-kb, low to ignore: 0.0000001), the gap between adjacent ROHs (--homozyg-gap, high to ignore: 5000 kcM or kb) and the window threshold to call a ROH (--homozyg-window-threshold, 0.0005) as predefined parameters in all calls. The sliding window in SNPs parameter (--homozyg-window-snp) and the maximum missing SNP allowance (--homozyg-window-missing), were also predefinde, but varied according to the minimum SNPs count (equal to --homozyg-snp, and --homozyg-snp × 0.05, respectively). The heterozygote allowance (--homozyg-window-het, 0 or 1) and the minimum SNP count to call a ROH (--homozyg-snp, 15, 25, 50, 75, 100, 150, 200, 300, 400) varied from call to call. Subjects with suspected uniparental disomy (at least 1 chromosome covered with >70% by ROH) and those with an extreme inbreeding (F_ROH_ ≥8%) were excluded after ROH calling, and prior to correlation coefficient calculation. ROH segments overlapping UCSC gap regions (centromeres, telomeres, contigs, …) were excluded.

We estimated the correlation between parental genetic relatedness offspring ROH length by using Spearman rank-order correlation. This approach was used for each of the sub-cohorts and for the 108 different combinations of ROH calling parameters. The parameters leading to the highest (positive) R were used for ROH calling in all triad members.

We attempted to replicate the findings by using genetic data in mothers and fathers. Given offspring genotype data was not available in these additional batches, we used predefined ROH calling parameters, based on the parameters obtained using our approach. These were the ROH calling parameters employed for the replication sub-cohorts, using genetic distance and with moderate pruning (R^2^> 0.5):

- --homozyg-window-snp 125 --homozyg-snp 125 --homozyg-kb 0.0000001 --homozyg-gap 5000 --homozyg-window-missing 125 * 0.05 --homozyg-window-threshold 0.0005 --homozyg-window-het 0 --homozyg-density 5000

For completeness, we compared our approach with widely used ROH calling parameters previously proposed by Joshi (9). The following parameters were employed in non-pruned data:

- --homozyg --homozyg-window-snp 50 --homozyg-snp 50 --homozyg-kb 1500 --homozyg-gap 1000 --homozyg-density 50 --homozyg-window-missing 5 --homozyg-window-het 1

As for the optimized parameters, we assessed the correlation between parental genetic relatedness and offspring ROH using Joshi’s parameters. Finally, we evaluated the overlap between autozygous segments called using the optimized and Joshi’s parameters. We matched autozygous segments for each individual with at least one autozygous segment detected using the optimized parameters, with autozygous segments detected using Joshi’s parameters. For each individual, we estimated the proportion of overlapping segments divided for the total segment length using the optimized parameters.

### Autozygosity calling in parent-offspring trios

The sub-cohort-specific parameters maximizing the coefficient of determination between offspring ROH length and parental genetic relatedness were used to call ROHs in all triad members. We calculated F_ROH_ for each subject as the sum of total ROH length divided by the total mappable distance (either in cM or bp) (15).

There is no single best measure of autozygosity, but we chose F_ROH_ as our primary measure and investigated the effects of other measures: excess homozygosity (F_HOM_), the total number of segments (NSEG) and the average length of ROH. F_HOM_ was calculated on a SNP-by-SNP basis using PLINK --het flag. Additionally, we estimated the time to the most recent common ancestor in generations as d/2k, where d is the genetic distance (100 cM) and k the individual average ROH length (58).

### Autozygosity mapping

After identifying ROH segments in each triad member of each sub-cohort, we split segments into unique intersecting segments in all sub-cohorts. Segments shared across sub-cohorts or unique to a single sub-cohort were kept (i.e., no other segment, from any cohort, had its start or end position included in any other segment). These segments were transformed into a matrix of binary calls, indicating whether an individual is autozygous for that particular segment. Only segments shared by at least 20 subjects (1 in 1250 or ∼0.08%) were kept.

Splitting segments into multiple non-overlapping segments means that additional correlation between close segments (some segments differed only in one subject) was introduced. The use of a Bonferroni p-value threshold correction would be overly conservative. To avoid this, we estimated the effective number of segments to identify segments with suggestive evidence. For each member of the triad, we ran principal components analysis on the segment matrix from each chromosome. The number of first eigenvalues explaining 0.995 of the variance in each chromosome was summed to obtain an effective number of segments. We identified high confidence segments using a Bonferroni corrected threshold of 0.05 / total number of autozygous segments.

Subsequently, low confidence segments were defined as segments surviving a Bonferroni correction of 0.05/ effective number of autozygous segments and were not high confidence segments; remaining segments were considered of no confidence. We created clumps of high and low confidence segments not farther from 0.5 cM from each other’s central position, and chose the segment with the lowest p-value within each clump as the top independent segment.

While we did not expect an effect from the paternal autozygous segments, we still ran survival models in fathers and used the results as a negative control.

### Long segment gene burden analysis

We conducted a gene burden analysis of long segments in all triad members using the same methods as for autozygosity mapping. For this analysis, we classified segment length according to Pemberton et al. (59). Briefly, we modeled the segment length distribution as a mixture of three Gaussian distributions: short segments may reflect ancient haplotypes, intermediate segments may have arised from background relatedness, and long segments may result from recent parental relatedness. We classified segment length using *Mclust* from the *mclust* package for each cohort. Given that our segment length was already large, we used segments classified as intermediate or long length for the burden test. Boundaries between different autozygous segment sizes for each sample and sub-cohort can be viewed in S7 Table. For each protein coding gene, we encoded gene burden as a binary variable indicating whether an individual had an autozygous segment partially or fully overlapping the gene transcription start and end (University of California Santa Cruz Table browser (60)). Whenever an autozygous segment overlapped several gene regions, each gene was used in the analysis.

### Survival analysis of imputed genetic data under a recessive model

Genotype data from all participants included in this study was pre-phased using Shapeit v2.790 (61) and then imputed at the Sanger Imputation Server using the Haplotype Reference Consortium v1.1 reference panel (62). Single genetic variant association analysis using hard called genotypes was performed using a recessive model. We fit separate models for mothers, fathers, and offspring for all genetic variants with an INFO score >0.4 that lay within the high or low confidence segments. We did not restrict minor allele frequency. All models were adjusted for sub-cohort, parity and the ten first principal components. Genetic variants surviving a genome-wide Bonferroni correction threshold (p-value< 5 × 10^−8^) were considered to be associated with time-to-spontaneous delivery and were subsequently tested for replication. A replication stage was performed using the same methods as for the discovery stage.

### Overall analysis strategy and rationale

Multiple analytical methods exist to model time-to-event data, which include, but are not limited to, Kaplan-Meier analysis, Cox proportional hazards regression and accelerated failure time models. While the Cox proportional hazards regression is the predominant method employed to model time-to-event data, we chose to use an accelerated failure time (AFT) model as an alternative approach. Both being multivariate survival analysis methods, we opted for AFT models over Cox proportional hazards regression for several reasons. Time-varying effects contribute to explain the heritability of gestational duration (63). Owing to the number of tests performed, we considered it unreasonable to assume a constant effect of ROHs on time-to-spontaneous delivery. Deviations from the proportional hazards assumption would lead to improper fitting of models and misleading inferences. AFT models have a more straightforward interpretation compared to proportional hazards. The covariates accelerate or decelerate survival time (64), affecting the rate at which an individual proceeds to the event. The coefficients can be directly interpreted as expanding (positive) or contracting (negative) median survival time - or that of any other percentile. Let S_F_(t) and S_R_(t) be the survival functions for individuals free and affected by ROH at a specific region. The AFT model specifies that

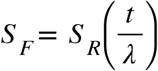

for any value of the survival time *t*, where *λ* is the acceleration failure rate, which reflects the impact of ROH presence on the baseline survival time.

AFT models require the specification of a probability density distribution for log(*t*). With the intention to obtain the probability density distribution that best fitted our data, we fitted an AFT model (R “flexsurv” package) with an intercept using five different distributions (exponential, Weibull, gamma, log-normal and log-logistic) on spontaneous delivery risk, using all pregnancies included in the MoBa cohort (n= 79155, events= 63169) adopting the same exclusion criteria as above-mentioned (except for recent white European ancestry and close genetic relatedness). Weibull distribution was the most appropriate probability density distribution based on the lowest Akaike information criterion (S8 Table). This probability distribution was used in all subsequent AFT models.

We estimated the effect of F_ROH_, F_HOM_, the total number of segments (NSEG), the average segment size, autozygosity mapping and gene burden on spontaneous onset of delivery risk in mothers, fathers and offspring in separate models using R “survival” package. The time scale was days until delivery, and the event, a spontaneous onset of delivery; pregnancies with deliveries initiated by induction or a planned cesarean section were censored (see Phenotype and covariates section for a detailed definition of spontaneous onset of delivery). Throughout the manuscript we report an estimate of the percent difference in time-to-spontaneous delivery (i.e. (exp(beta) - 1) × 100) for any survival time quantile. We ran a total of two models for both F_ROH_, F_HOM_, NSEG, the average length of segments and time to most common ancestor for each triad member group: a crude model adjusting for sub-cohort and model 1 adjusting for sub-cohort, parity (nulliparous vs. multiparous), 10 principal components, maternal educational attainment and household income. Thus, we used a Bonferroni corrected significance threshold by accounting for the number of exposures in each triad member, and considered significant if p-value was below 0.01 (0.05 / 5 autozygosity measures). Autozygosity mapping and gene burden models were adjusted for sub-cohort, F_ROH_, parity and the first ten principal components.

Code for data manipulation and analysis was structured using Snakemake (65), and is available at https://github.com/PerinatalLab/ROH.

### Segment and genetic variants annotation

All segments and genetic variants were mapped to the nearest protein-coding gene. Gene transcription coordinates were downloaded from the University of California Santa Cruz Table browser (60). Functional data of genetic variants using a recessive model, the expected mode of action in variants within our high confidence segments, is scarce. Thus, we mapped the identified genes with genes known to affect phenotypes with a recessive inheritance from the Online Mendelian Inheritance in Man (OMIM) data (66). We annotated variants within high confidence segments with moderate or high impact (according to Ensembl VEP (67)) using gnomAD version 2.0.1 (68).

## Data Availability

Access to genotypes and phenotypes can be obtained by direct request to the Norwegian Institute of Public Health (https://www.fhi.no/en/studies/moba/for-forskere-artikler/gwas-data-from-moba/).

## Ethics statement

This study was approved by the Regional Committee for Medical and Health Research Ethics from Norway (2009/1387).

## Acknowledgments

The Norwegian Mother, Father, and Child Cohort Study is supported by the Norwegian Ministry of Health and Care Services and the Ministry of Education and Research, NIH/NIEHS (contract no N01-ES-75558), NIH/NINDS (grant no.1 UO1 NS 047537-01 and grant no.2 UO1 NS 047537-06A1). This work is supported by a grant from the Burroughs Wellcome Fund (10172896). We thank the Norwegian Institute of Public Health (NIPH) for generating high-quality genomic data. This research is part of the HARVEST collaboration, supported by the Research Council of Norway (#229624). We also thank the NORMENT Centre for providing genotype data, funded by the Research Council of Norway (#223273), South East Norway Health Authority and KG Jebsen Stiftelsen. We further thank the Center for Diabetes Research, the University of Bergen for providing genotype data and performing quality control and imputation of the data funded by the ERC AdG project SELECTionPREDISPOSED, Stiftelsen Kristian Gerhard Jebsen, Trond Mohn Foundation, the Research Council of Norway, the Novo Nordisk Foundation, the University of Bergen, and the Western Norway health Authorities (Helse Vest). LJM and GZ were supported by the March of Dimes Prematurity Research Center Ohio Collaborative. We are grateful to all the participating families in Norway who take part in this ongoing cohort study.

## Notes

### Competing Interest Statement

The authors have declared no competing interest.

### Author Declarations

Regional Committee for Medical and Health Research Ethics from Norway (2009/1387)

